# Liver histopathology in COVID 19 infection is suggestive of vascular alteration

**DOI:** 10.1101/2020.05.06.20092718

**Authors:** Aurelio Sonzogni, Giulia Previtali, Michela Seghezzi, Maria Grazia Alessio, Andrea Gianatti, Lisa Licini, Pietro Zerbi, Luca Carsana, Roberta Rossi, Eleonora Lauri, Alessandro Pellegrinelli, Manuela Nebuloni

## Abstract

COVID-19 breakout in Italy has caused a huge number of severely ill patients with a serious increase in mortality. Although lungs seem to be the main target of the infection very few information are available about liver involvement in COVID-19 infection, that could possibly evocate a systemic disease targeting a lot of organs. Since now there are no reports of large series of histological evaluation of liver morphology in this setting. Knowledge of histological liver findings connected to clinical data is crucial in management of this disease. Post-mortem wedge liver biopsies from 48 patients died for COVID-19 infection were available from two main hospitals located in northern Italy, Lombardy; all sample were obtained during autopsies. No patient has a significant clinical complain of liver disease or signs of liver failure before and during hospitalization; for each of them laboratory data focused on liver were available. All liver samples showed minimal inflammation features; on the other side, many histological pictures compatible with vascular alterations were observed, characterized by portal vein braches number increase associated with lumen massive dilatation, partial or complete recent luminal thrombosis of portal and sinusoidal vessels, fibrosis of portal tract, focally severely enlarged and fibrotic. Our preliminary results concerning histological liver involvement in COVID-19 infection confirm the clinical impression that liver failure is not a main concern and this organ is not the target of significant inflammatory damage; histopatological findings are highly suggestive for marked alteration of intrahepatic blood vessel network secondary to systemic alterations induced by virus that could target, besides lung parenchyma, cardiovascular system, coagulation cascade or endothelial layer of blood vessels.

## Introduction

The novel coronavirus infection (named COVID-19) has been first diagnosed in December 2019 in Wuhan, China, and then it has spread all around the world. On 11th March 2020, World Health Organization declare a worldwide pandemic status. The majority of the patients develop a mild self-resolving infection, but some cases present pneumonia and the most severe of them progress to a systemic disease with multiple organ dysfunction.

Bergamo, a city located in northern Italy 40 miles far from Milan, became quickly the epicenter of the Italian COVID-19 outbreak: until now more than 9000 people are infected and at least 2500 persons died. Diagnosis for COVID-19 infection was performed by PCR reaction on nose-pharyngeal swab.

Bergamo area count about 900000 inhabitants and Papa Giovanni XXIII is the main city hospital, hosting about 900 beds with 90 intensive care unit beds but, during this emergency situation, 80 ICU workstation were completely devoted to the most severe COVID-19 patients. Even the area of Milan registered a high number of infected patients and at L. Sacco Hospital, a referral third level institution in Milan focused on infectious diseases care, hosting 525 beds

Since the outbreak of COVID-19 pandemia on February 21^st^, 900 patients were admitted (131 in intensive care unit; 140 treated by CPAP support) in Bergamo Hospital; medium age was 61 ± 11,5 (median 62) years. During the same period, 230 COVID-19 patient died between February 21^st^ and March 25^th^ in Bergamo Hospital.

To our knowledge, this is the first time a large series of liver histopathology findings are described for COVID-19 patients died for respiratory failure; significant liver clinical involvement has never been observed in such type of patients and neither liver pathology in COVID-19 infection nor clinical-pathological correlations have been reported.

## Methods

Wedge liver sample were obtained post-mortem in 48 COVID-19 positive patients ( 35 males, 13 females; medium age 71,2 years; range 87-32 years), all of them deceased for severe respiratory failure after a variable duration of hospitalization, ranging from 1 to 21 days (median duration 7 days); sampling procedure was partial autopsy limited to lungs, heart and liver in 30 patients and a complete autopsy in 18 cases excluding brain. No patient had overt previous history of liver disease or developed clinical signs or symptoms of liver failure during hospital stay. 33 patients reported significant one or more comorbidity conditions (morbid obesity with BMI >30, chronic heart failure, hypertension).

The autopsies were performed in Airborne Infection Isolation autopsy rooms and the personnel used the correct Personal Protection Equipment (PPE), according to "Engineering control and PPE recommendations for autopsies". Only skilled pathologists were enrolled to perform post-mortem procedures.

A medium of two tissue blocks were taken random from each liver as macroscopic aspect was normal; the size of all the blocks obtained were comparable. Tissues were fixed in 10% buffered formalin for >48 hours and embedded in paraffin. Three-mm paraffin sections were stained with by hematoxylin and eosin Histological exam of other organs sampled during autopsies is still pending due to complexity of lesions found microscopically mainly in lungs.

Each wedge liver sample contained at least 20 portal fields; histological examination was performed blindly by experienced pathologists confident with liver non neoplastic histopathology. In few cases additional immunohistochemical staining for inflammatory cells (CD3 and CD20) or for easier detection of endothelial layer and vessel conformation (CD34, factor VIII) was obtained. Laboratory findings were provided after histological examination and are summarized in table 1;

Not all patients undergoing post-mortem liver biopsy got the same laboratory profiling, due to different duration of hospitalization. Based on published literature, the development of coagulopathy is a poor prognostic features for COVID-19 patients [1-3], so when available, the highest results for coagulation and liver function tests were collected during the hospital stay: prothrombin time (PT, expressed as international normalized ratio -INR-), D-dimer and platelet count; aspartate transaminase (AST), alanine transaminase (ALT) and gamma-glutamyl transpeptidase (GGT). To compare and evaluate values for AST, ALT and GGT determination, results are expressed as ratio with the upper reference levels (fold-xULR).

## Results

Main liver histological findings obtained by microscopic examination are detailed reported in table 2. Lobular architecture was well preserved in all samples; focal inflammatory infiltrate (mainly made by scattered portal and lobular lymphocytes, mainly CD4 T lymphocytes) was detected and only mild/moderate portal fibrosis was histologically proved.

In spite of expected inflammatory damage, the clearly emerging morphological features were characterized by diffuse alterations of intrahepatic blood vessels structure (portal branches and sinusoids) and variable degree of partial and complete luminal recent thrombosis, features very similar to those described in patients affected by congenital deficiency of coagulation factors or in clinical conditions characterized by impaired liver blood circulation such as hepato-portal syndrome. Majority of portal fields showed an increase in number of portal veins associated with luminal severe dilatation; in a large number of sample lumen focally herniated was detected in peri-portal liver parenchyma. Kupffer cells were found extremely activated with large cytoplasm containing necrotic debris. Steatosis was diagnosed in a large number of samples, but this finding was largely forecast, as a relevant number of patients was obese.

Main histological features, present with various degree in all specimens, were very suggestive for hepatic vascular involvement, acute (thrombosis, luminal ectasia) and chronic (fibrous thickening of vascular wall or phlebosclerosis, abnormal asset of portal intrahepatic system) (table 1). Moreover CD 34 (endothelial marker) performed in most of the biopsies, besides confirm an abnormal configuration of intrahepatic blood vessels, was able to decorate a peri-portal network of sinusoidal vessels; this finding is totally abnormal and is usually reported as sinusoidal arterialization, due to an increase pressure of blood (table 1). Immunohistochemical search for C4d, performed in all samples, was completely negative.

The evaluation of liver function tests showed that all the patients, but one, have an alteration of at least one of the three parameters taken into consideration, the positivity rate was as follow: AST 40/41 (97.6%), ALT 25/41 (61.0%), GGT 20/26 (76.9%). Using a cut off of 500 ng/dL, 25/26 (96.1%) patients had high D-dimer values; the distribution of PT-INR values and platelet count, along with the previous described results, are summarized in figure 3.

**Table 1.** Main histopathological findings in liver wedge biopsies

|  | **All patients**<br>**n= 48** |
| --- | --- |
| **Portal vein phlebosclerosis** |  |
| absent | 0 |
| focal (up to 25% of the portal tracts) | 14 |
| multifocal (25-75% of the portal tracts) | 11 |
| diffuse (>75% of the portal tracts) | 4 |
| **Herniated portal vein (Aberrant portal vessels),** |  |
| absent | 12 |
| focal (up to 25% of the portal tracts) | 18 |
| multifocal (25-75% of the portal tracts) | 13 |
| diffuse (>75% of the portal tracts) | 5 |
| **Type of Herniated portal vein** |  |
| connected to sinusoids | 47 |
| connected to hepatic veins | 2 |
| **Periportal abnormal vessels** |  |
| focal (up to 25% of the portal tracts /parenchyma) | 27 |
| multifocal (25-75% of the portal tracts /parenchyma) | 18 |
| diffuse (>75% of the portal tracts /parenchyma) | 3 |
| **Fibrosis** |  |
| portal fibrosis | 29 |
| incomplete fibrous septa | 8 |
| **Lobular inflammation** |  |
| Absent | 24 |
| Mild | 23 |
| Moderate | 1 |
| Severe | 0 |
| **Portal inflammation** |  |
| Absent | 16 |
| Mild | 32 |
| Moderate | 0 |
| Severe | 0 |
| **Vascular thrombosis** |  |
| Partial portal | 24 |
| Complete portal | 11 |
| Incomplete sinusoidal | 7 |
| Complete sinusoidal | 6 |
| **Steatosis** |  |
| Microvesicular (% of affected hepatocytes) | 3(5,5,20%) |
| Macrovesicular (%) | 1(20%) |
| Mixed (%) | 22<br>(50,50,50,40,20,10,30,10,10,10,20,50,10,20,30<br>25, 30, 10, 20, 60, 20,15%) |

**Figure 1.**
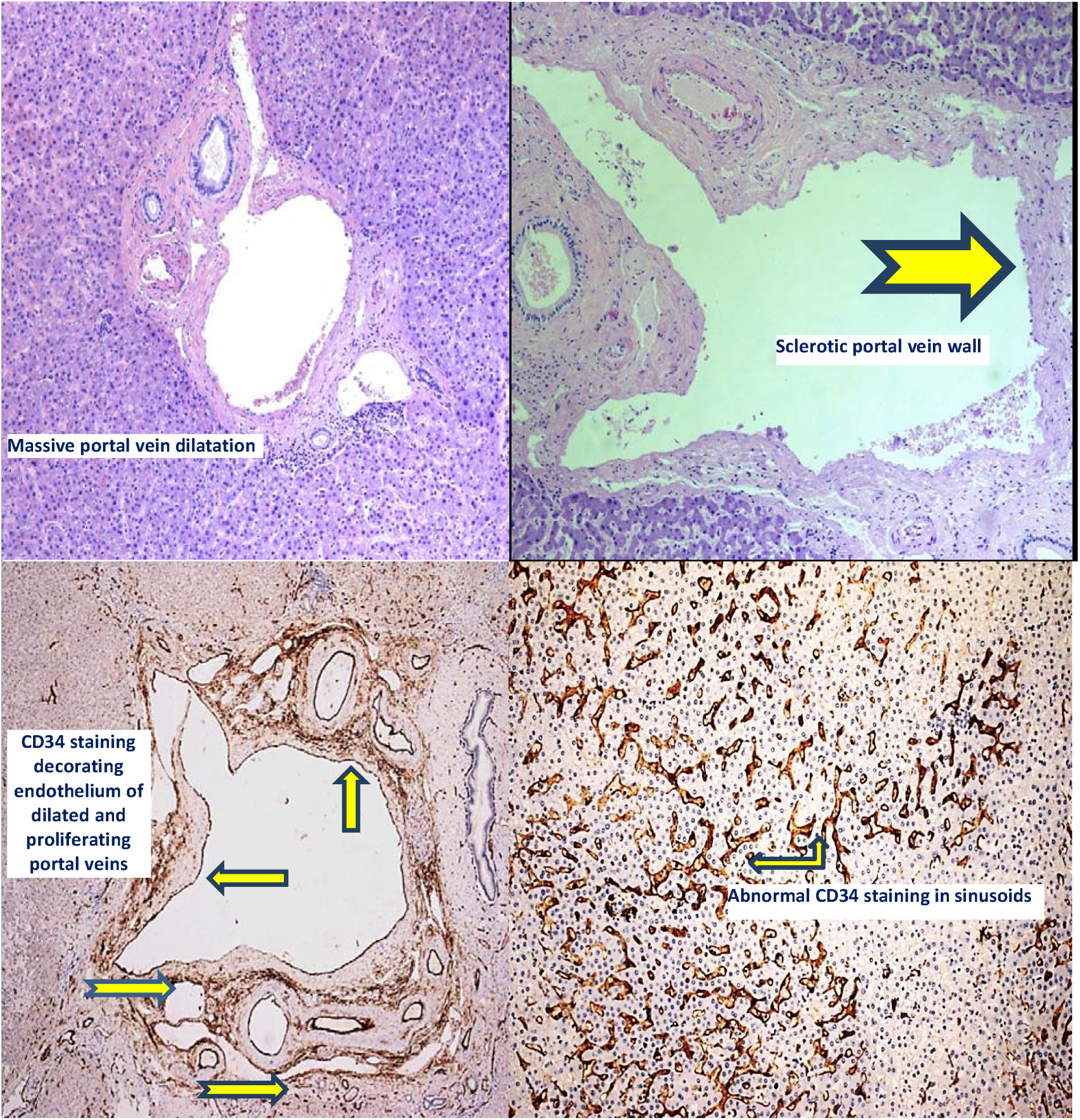
Abnormal structure of intrahepatic portal tree characterized by number increase, severe and diffuse lumen dilatation (upper left) (H&E, 400X); dilated vessels also show sclerotic and thickened wall (upper right) (HeE, 400X). Immunohistochemical staining for endothelium (CD34) decorated in brown a severe dilated portal vein (bottom left) and many sinusoids, unstained in normal conditions (bottom right) (400X).

**Figure 2.**
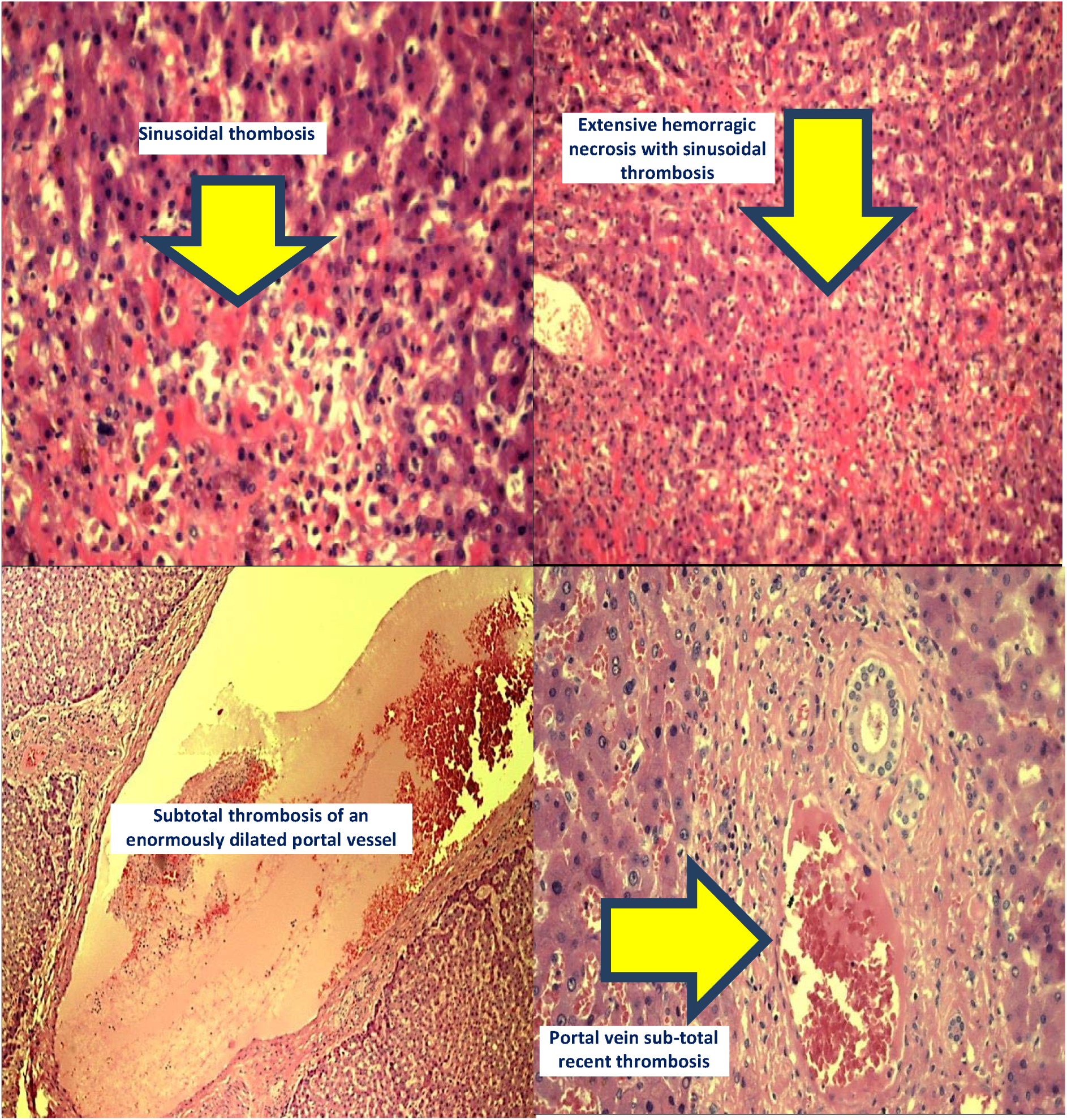
Diffuse thrombotic phenomena in liver intra-parenchymal vessels; sinusoidal involvement is characterized by intravascular fresh clots (upper left) (H&E; 600X) causing liver cells hemorragic necrosis (upper right) (H&E; 400X); portal thrombosis may be present in large (bottom left) and in terminal branches (bottom right) (H&E; 600X).

**Figure 3.**
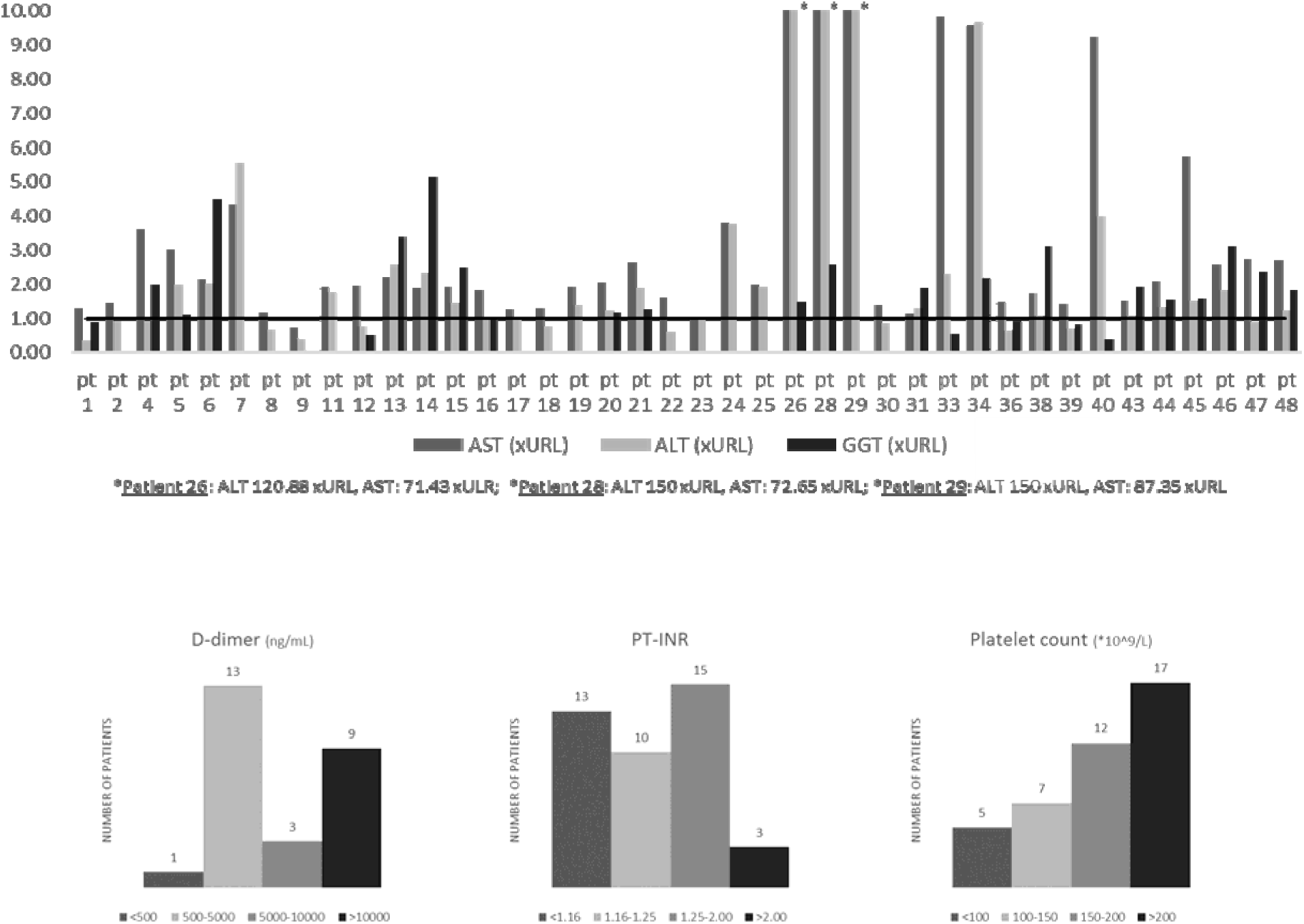
Main liver function tests, clotting main parameters and tests of thrombo-embolic condition; some data are not available for few patients due to a very short time of hospital stay. (AST: Aspartate Transaminase; ALT: Alanine Transaminase; GGT: Gamma-Glutamyl Transpeptidase; PT-INR: Prothrombin Time-International Normalized Ratio; URL: Upper Reference Limit)

## Discussion

Morphological studies concerning description and interpretation of liver parenchymal alterations induced or related to COVID-19 infection are completely lacking; till now only a very weak post-mortem report is available in Chinese language and focused mainly on lung destructing lesions [4]. Knowledge of liver pathology is totally blank and there are no persuasive correlations available at present time among clinical features, laboratory findings and liver histology.

Since outbreak of a severe pandemic in Lombardy, Italy, we got the opportunity to examine a large liver samples from wedge post-mortem biopsies; moreover, a lot of laboratory test for these patients were available, thus allowing a correlation between morphology and clinical findings. Liver histology examination performed on these samples demonstrated that this organ is not involved in any significant inflammatory response to COVID-19 infection and our data strongly support the first clinical impression that liver is not a consistent concern in management of COVID-19 affected patients.

All our morphological findings obtained in deceased COVID-19 positive patients are otherwise consistent with vascular related damage due to impaired blood flow, generating lesions similar to the histological picture observed in hepato-pulmonary syndrome and in obliterative portal venopathy [5,6]; a tentative physio-pathological explanation could be based on an increased blood flow within the liver, sometimes related also to heart distress, or/and thrombotic phenomena in portal and sinusoidal vessels, modifying intrahepatic blood circulation; in addition some clinical details and laboratory findings point in this peculiar direction (i.e. high D-dimer levels in blood) associated with our unpublished findings in lungs affected by COVID-19 infection, showing an huge number of thrombotic medium and small caliber branches of pulmonary arteries. To support this hypothesis, we underline that some cases of obliterative portal venopathy described by Guido and al. [7] were associated with congenital or acquired anomalies of some factors activated during clotting cascade; another putative mechanism could be an immunological attack to endothelial layer. Our histological findings strengthen the hypothesis that the derailment of coagulation process or impairment of blood circulation or endothelial damage could be main triggers mechanism in pathogenesis of COVID-19 damage not only within the liver but possibly also in many other solid organs. This theory fits really well with some recently published observations by a British group [8] based on pure clinical ground, focusing the main rule of clotting system in COVID-19 liver damage.

Moreover, the abnormal laboratory findings pointing to a necrotic process targeting liver cells and inducing high levels of transaminases in some patients should be explained by vascular portal and sinusoidal thrombosis, leading to focal parenchymal necrosis and liver cells accelerated apoptosis.

A further very crucial question is if any previous chronic asymptomatic condition such phlebosclerosis noted in all biopsies although in variable degree of severity, could be a hidden predisposing condition to liver damage; this peculiar feature could be related to long standing sub-clinical process partially related to the advanced age of the patients. Further and deeper studies on liver samples are obviously ongoing to get quickly more detailed and clinically relevant information. A promising field of great interest should be the study of endothelial cells alterations induced by virus infection and interactions with clotting process.

## Data Availability

Available to all

## Notes

None of the authors declare a conflict of interest

This work has no financial support

### Competing Interest Statement

The authors have declared no competing interest.

### Clinical Trial

None

### Funding Statement

No funding support

